# DEFINING THE DIVERSITY OF HNRNPA1 MUTATIONS IN CLINICAL PHENOTYPE AND PATHOMECHANISM

**DOI:** 10.1101/2021.02.02.21250330

**Authors:** D Beijer, HJ Kim, L Guo, K O’Donovan, I Mademan, T Deconinck, K Van Schil, CM Fare, LE Drake, AF Ford, A Kochański, D Kabzińska, N Dubuisson, P Van den Bergh, NC Voermans, RJLF Lemmers, SM van der Maarel, D Bonner, JB Sampson, MT Wheeler, A Mehrabyan, S Palmer, P De Jonghe, J Shorter, JP Taylor, J Baets

## Abstract

Mutations in *HNRNPA1* encoding heterogeneous nuclear ribonucleoprotein (hnRNP) A1 are a rare cause of amyotrophic lateral sclerosis (ALS) and multisystem proteinopathy (MSP). hnRNPA1 is part of the group of RNA-binding proteins (RBPs) that assemble with RNA to form ribonucleoproteins. hnRNPs are a major subclass of evolutionarily conserved RBPs that are primarily concentrated in the nucleus and are heavily involved in pre-mRNA splicing, mRNA stability and transcriptional/translational regulation. During times of stress, standard translational programming is interrupted, and hnRNPs, mRNA, and other RBPs condense in the cytoplasm, forming liquid-liquid phase separated (LLPS) membraneless organelles termed stress granules (SGs). SGs are central to the pathogenesis of (neuro-)degenerative diseases, including ALS and inclusion body myopathy (IBM). hnRNPs and other RBPs are critical components of SGs. Indeed, the link between SGs, hnRNPs, and neurodegenerative diseases has been established by the identification of additional mutations in RBPs that affect SG biology, including FUS, TDP-43, hnRNPA1, hnRNPA2B1, and TIA1, each of which can directly lead to ALS, IBM and other related neurodegenerative diseases. Here, we report and characterize four novel *HNRNPA1* mutations and two known *HNRNPA1* mutations, previously reported as being causal for ALS, in a broad spectrum of patients with hereditary motor neuropathy (HMN), ALS, and myopathy. Our results show the different effects of mutations on hnRNPA1 fibrillization, liquid-liquid phase separation, and SG dynamics, indicating the possibility of different underlying pathomechanisms for *HNRNPA1* mutations with a possible link to the clinical phenotypes.

## INTRODUCTION

Heterogeneous nuclear ribonucleoprotein A1 (hnRNPA1) is a member of a large class of RNA-binding proteins (RBPs) that assemble with RNA to form ribonucleoproteins (RNPs). Heterogeneous nuclear ribonucleoproteins (hnRNPs) are a major subclass of evolutionarily conserved RNPs that are primarily concentrated in the nucleus and are heavily involved in pre-mRNA splicing, mRNA stability, miRNA maturation, transcriptional regulation, translational regulation and telomere biogenesis [53,47,70,19,13,36,34]. Over the last decade, the link between hnRNPs and neurodegenerative diseases has been established by the identification of mutations impacting several RBPs, including FUS, TDP-43, hnRNPA1, hnRNPA2B1, matrin-3 (MATR3), and TIA1 as causative for amyotrophic lateral sclerosis (ALS), frontotemporal dementia (FTD), and hereditary inclusion body myopathy (hIBM) phenotypes. Missense mutations in the prion-like domain (PrLD) of hnRNPA1 (p.D262V/N and p.N267S) have been identified as a cause of ALS [MIM: 615426] and multisystem proteinopathy 3 (MSP3) [MIM: 615424] [32,3]. Subsequent confirmation of ALS-causing mutations in hnRNPA1 were found to also impact the PrLD (p.G264R) [29]or, alternatively, the nuclear localization sequence (NLS; p.288S/A) [42,51]. Interestingly, the p.D262N mutation in *HNRNPA1* was also reported in two IBM families with a pure muscular phenotype, illustrating the expressivity differences that frequently arise from HNRNP mutations. Several modest-sized genetic screenings by different research groups failed to identify a significant number of additional families with *HNRNPA1* mutations in a total of 2485 patients with ALS, frontotemporal dementia (FTD), MSP, hereditary inclusion body myopathy (hIBM) or other myopathies, suggesting that disease-causing mutations in *HNRNPA1* may be relatively rare [6,62,35,60,31]. Recent work has identified stress granules (SGs) as being at the nexus of RBP pathology and neurodegenerative diseases [4,5,21,39,43,48]. SGs are liquid-liquid phase separated cytoplasmic membraneless organelles containing proteins from a variety of different RNA processing and translation pathways, including a variety of RBPs that are strongly correlated with neurodegeneration [21]. SGs are mostly encompassing mRNA transcripts, with approximately 80% of the SG consisting of mRNAs from essentially every expressed gene [50,30]. SGs also specifically contain small ribosomal subunits; translation initiation factors (eIF3, eIF4E, eIF4G); and hnRNPs and other RBPs, such as TIA-1, HuR, PABP, and TTP. SGs form following stress exposure and mature from numerous small inclusions that fuse over time. Importantly, SGs are dissolved once stress is diminished, allowing the component molecules to return to their soluble function [28]. As a subcellular compartment, SGs are central to the pathogenesis of degenerative diseases, as disease-causing mutations in RBPs are associated with accumulation of persistent SGs in ALS, FTD, and IBM [32,39,55,5,22].

Interestingly, neurodegenerative diseases are often characterized by pathological proteinaceous inclusions, a subset of which contain select SG markers. A good example is deposition of TDP-43, which is a prominent feature in large portions of familial and sporadic ALS cases, as well as nearly all cases of sporadic IBM and hIBM [32,33,66,27,43]. The formation of these pathological inclusions may be due to dysregulation of the SG response, most prominently driven by the failure of SGs to disassemble. As such, it is critical to distinguish if the inclusions observed in *post mortem* tissues are broadly composed of SG proteins, if SG proteins are themselves recruited to pre-formed inclusions, or both [4]. Thus far, we are limited to findings that show RBP depletion from the nucleus and deposition in cytoplasmic aggregated structures to be a consistent pathology, for which the underlying mechanisms of *HNRNPA1* and other RBP deposition remains largely unknown.

Recent work on the broader group of ventral horn, peripheral nerve and muscle diseases –including inherited peripheral neuropathy (IPN), hereditary motor neuropathy (HMN) and ALS – have demonstrated a significant genetic overlap, including both pleiotropy and genetic heterogeneity. This overlap is aptly illustrated by findings where causative genes do not exclusively give rise to the phenotype for which they were initially described, as is apparent for mutations in *SETX, DCTN1* and *SIGMAR1*, which are causative for both ALS and HMN [1,49,7].

In this study, we present patients with four novel *HNRNPA1* mutations and two known *HNRNPA1* mutations. These mutations expand the genetic and clinical spectrum of neurodegenerative diseases by demonstrating their involvement in complex IPN, atypical ALS and myopathy phenotypes. In addition, we present results that show the different effects of mutations on hnRNPA1 fibrillization, liquid-liquid phase separation (LLPS), and SG dynamics, indicating the possibility of alternative underlying pathomechanisms for *HNRNPA1* mutations with a possible link to the clinical phenotypes.

## METHODS

### Study participants

We describe a total of six families: three families with an isolated patient, and three families with a dominant inheritance of motor neuron or muscle disorders (Table 1). Samples from these six families were subjected to next-generation sequencing. In addition, a total of 547 patients diagnosed with dHMN, CMT2, intermediate CMT, spinal muscular atrophy (SMA), ALS, hereditary spastic paraplegia (HSP), or myopathy were screened for *HNRNPA1* mutations. For all patients, genomic DNA was extracted from blood samples obtained from patients and family members using standard methods. All patients or their legal representatives signed informed consent before enrollment. The ethical review board of the University of Antwerp approved this study.

**Table 1.** *HNRNPA1* mutations leading to a broad spectrum of phenotypes.

| Individual | A:II:1 | B:II:1 | B:II:2 | C:II:2 | C:III:1 | D:II:1 | E:II:2 | F:I:1 |
| --- | --- | --- | --- | --- | --- | --- | --- | --- |
| Gender | Male | Female | Female | Female | Male | Female | Male | Male |
| Parental consanguinity | No | Yes | Yes | No | No | No | No | No |
| Current age range (age at death) | 40-50 years | 40-50 years | 40-50 years | 40-50 years | 10-20 years | 30-40 years | 40-50 years | † 60-70 years |
| **Mutation** |  |  |  |  |  |  |  |  |
| Genomic position (hg19) | Chr12: 54678040 A>G | Chr12: 54677706 C>G | Chr12: 54677706 C>G | Chr12: 54678095 T>G | Chr12: 54678095 T>G | Chr12:54677766-54678265 deletion 500bp deletion | Chr12:54677629A >T | Chr12:54678095 T>C |
| cDNA | NM_002136: c.908-2A>G | NM_002136: c.862C>G | NM_002136: c.862C>G | NM_002136: c.961T>G | NM_002136: c.961T>G | NM_002136: c.907+15_*5-68del | NM_002136: c.785A>T | NM_002136: c.961T>C |
| Protein | p.G304Nfs*3 | p.P288A | p.P288A | p.*321Eext*6 | p.*321Eext*6 | p.G304Nfs*3 | p.D262V | p.*321Qext*6 |
| **Clinical features** |  |  |  |  |  |  |  |  |
| Overall phenotype | Distal pure motor neuropathy | Motor neuron disease | Motor Neuron disease | Distal myopathy | Distal myopathy | Distal myopathy | Distal myopathy | Distal myopathy |
| Decade of onset | 2nd | 3rd | 5th | 1st | 1st | 3rd | 4th | 2nd |
| Symptoms at onset | Wasting hand intrinsics | Paresis of left hand without sensory impairment | Swallowing difficulties and dysarthria | Difficulties with hand function, mild facial weakness, fatigue at age 12 | Foot drop R | Hand weakness while writing | Weakness foot R frequent tripping and falls | Wasting of hand intrinsics, weakness lower limbs |
| Age at last examination | 40-50 years | 40-50 years | 40-50 years | 40-50 years | 10-20 years | 30-40 years | 40-50 years | 60-70 years |
| Walking (canes, wheelchair,...) | Ambulant | Ambulant | Ambulant | Wheelchair use: intermittent (20-30 years) permanent (30-40 years) | Ambulant | Ambulant | Cane, hand rails when going up the stairs | Wheelchair bound |
| Walking on toes | Yes | Yes | Yes | No | Yes | No | Yes | No |
| Walking on heels | No | Yes | yes | No | No | No | No | No |
| Foot deformities | Slight *pes cavus* | - | No | Slight hammer toes | *Pes planus* on right foot | No | High arched feet, hammer toes | *Pes equinovari* |
| **Muscle atrophy UL** | Hand intrinsics, distal forearm | R>L | Hand intrinsics R | Generalized | Thenar L | Hand intrinsics R>L | No | Pronounced |
| **Muscle atrophy LL** | Yes | No | No | Generalized | No | Yes | No | Pronounced |
| **Weakness proximal UL** | Not measured | Deltoid >biceps and triceps +++ bilaterally, neck flexors and abdominals | No | Shoulder abduction 2/5, Shoulder anteflexion mild | No | No | No | Biceps 0/5, triceps 2-/5, shoulder abduction 1-3/5 |
| **Weakness distal UL** | Not measured | Hand intrinsics +++ bilaterally | Weakness (2/5) right abductor pollicis brevis and I interosseous | Hand intrinsics 2/5, finger flexion/extension 0/5 | Opponens pollicis MRC 5-/5 L | finger and wrist extension 1/5 R, 4-4+/5 L | No | 0/5 |
| **Weakness proximal LL** | Not measured | No | No | Knee extension 1/5, hip adduction/abduction 1/5 | No | No | Knee flexion/extension 4+/5 | 0/5 |
| **Weakness distal LL** | Not measured | No | No | 0/5 | Foot plantar flexion 5/5, Foot/toe dorsiflexion 3/5 L, 0/5 R | ankle dorsiflexion 0/5 R, 4-/5 L | Foot eversion 4+/5, inversion 5/5, dorsiflexion 3+/5, plantar flexion 4-/5 | tibialis anterior 2/5, otherwise 0/5 |
| **Reflexes UL** | Very brisk | Very brisk | Brisk | Absent | Normal | Trace | Normal | Absent |
| **Reflexes LL** | Brisk | Absent | Normal | Absent | Normal | Trace | Normal | Absent |
| **Predominance in the hands** | Yes | Yes but also severe in deltoids | Yes | Yes | No | Yes | No | Yes (initial presentation) |
| **Sensory involvement** | - | - | - | - | - | No | No | No |
| **Pyramidal tract signs (Babinski sign, clonus, ...)** | No | Bilateral Hoffmann-Trömner, no Babinski sign | Right Hoffmann-Trömner, no Babinski sign | No | No | No | No | No |
| **Respiratory difficulties** | No | Vital capacity 59% | No | Vital capacity 70%, aggravated during cold or fever | No | FVC 76% | No | Non-invasive ventilation (BiPAP), cough assist |
| **Facial weakness** | No | Yes | No | Yes, very pronounced | No | Yes | No | Yes, very pronounced |
| **Other clinical features** | Mild thoracic scoliosis, depression, mild deformation of the L1 and L2 vertebrae | - | Tongue atrophy and dysarthria | Dysarthria, most likely due to the very severe facial weakness | - | Asymmetry of weakness, scapular winging | - | Percutaneous endoscopic gastrostomy, muscle contractures UL and LL, dysphagia, titubation |
| **Additional testing** |  |  |  |  |  |  |  |  |
| **NCS** | Pure motor axonal neuropathy | Pure motor axonal neuropathy | - | Pure motor axonal neuropathy | Normal | Decreased amplitude median and peroneal motor | low amplitude peroneal motor consistent with chronic myopathy | Normal, very small CMAPs |
| **Needle EMG** | Chronic neurogenic | Not performed | Not performed | Myopathic, proximal UL | Myopathic, distal and proximal | Irritative distal predominant myopathy. | Chronic myopathic changes with muscle membrane irritability in LL | Chronic myopathic |
| **Nerve biopsy** | Very mild axonal neuropathy with secondary demyelination (20-30 years) | Not performed | not performed | Not performed | Not performed | Not performed | Not performed | Normal |
| **Muscle biopsy** | Not performed | Not performed | Not performed | Angular fibers, no type grouping (10-20 years) | Slight clustering type 1 fibers (0-10 years) | Myofibrillar disorganization, rimmed vacuoles no inflammation (30-40 years) | Chronic myopathy with rimmed vacuoles (40-50 years) | Type 1 atrophy, limited rimmed vacuoles, centralized nuclei type 2 fibers (10-20 years) |

L, left: LL, lower limb(s); MRC, medical research council; NCS, nerve conduction studies; R, right: UL, upper limb(s); (-), absent/unknown

### Next-generation sequencing

#### Family A

Whole-exome sequencing was performed on high molecular weight genomic DNA sample of A:II:1 (Fig. 1). The Nextera Rapid Capture Expanded Exome kit (62Mb) (Illumina) was used for exome enrichment, with subsequent library sequencing on a HiSeq 2500 Illumina platform. The Burrows‐Wheeler Aligner (BWA) tool was used to perform the sequence alignment to the reference genome (hg19, UCSC Genome Browser) [37]. Variant calling was performed with Genome Analysis Toolkit (GATK) Unified Genotyper, and for the annotation and filtering the Clinical Sequence Analyzer and Miner (Wuxi NextCODE) was used [45]. Filtering of the data occurred using the following criteria: no occurrence or a frequency ≤ 0.5% of the variants in public exome variant repositories (Exome Aggregation Consortium, 1000 Genomes Project, Exome Variant Server, in‐house data); variants with impact on the encoded protein (missense, nonsense, frame shift, in-frame insertions/deletions and splice site variants); read depth ≥ 7; minimal heterozygous call percentage ≥ 20%; minimal homozygous call percentage ≥ 66%. For possible pathogenic variants, Sanger sequencing was used for validation and segregation analysis in the patient and unaffected parents.

**Fig. 1.**
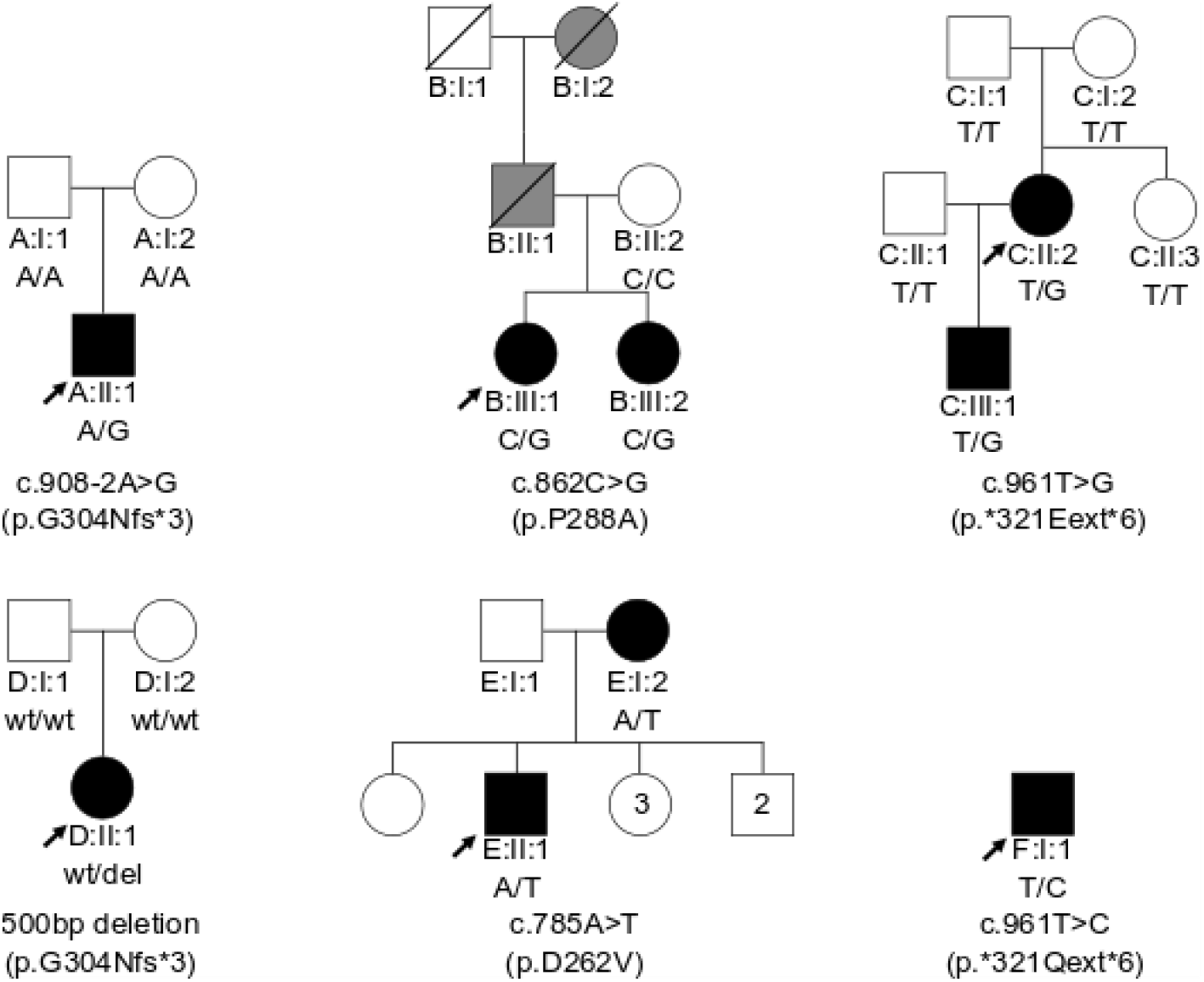
Heterozygous *HNRNPA1* mutations in six families. Pedigrees of families A, B, C, D, E and F with their respective mutation, and the segregation of each by genotype, showing affected (black), unaffected (white) and hearsay affected (grey) individuals. Arrows indicate probands.

#### Family B

Whole-exome sequencing was performed on high molecular weight genomic DNA samples of individuals B:II:1 and B:II:2 (Fig. 1). The SeqCap EZ Human Exome Library v3.0 (64Mb) (Roche) was used for exome enrichment, with subsequent library sequencing on a NextSeq 500 Illumina platform. The Burrows‐Wheeler Aligner (BWA) tool was used to perform the sequence alignment to the reference genome (hg19, UCSC Genome Browser) [37]. Variant calling was performed with Genome Analysis Toolkit (GATK) Unified Genotyper and SAMtools [45,38]. For the annotation and filtering we used the in‐house developed tool GenomeComb [56]. Filtering of the data occurred using the following criteria: no occurrence or a frequency ≤ 0.5% of the variants in public exome variant repositories (Exome Aggregation Consortium, 1000 Genomes Project, Exome Variant Server, in‐house data); variants with impact on the encoded protein (missense, nonsense, frame shift, in-frame insertions/deletions and splice site variants); read depth ≥ 7; minimal heterozygous call percentage ≥ 20%, minimal homozygous call percentage ≥ 66%. For possible pathogenic variants, Sanger sequencing was used for validation and segregation analysis in the index patient and a combination of three affected and unaffected family members.

#### Family C

Whole-genome sequencing was performed on high molecular weight genomic DNA samples of C:I:1, C:I:2, C:II:2 and C:III:1 (Fig. 1) using a HiSeq 2500 Illumina platform. The Burrows‐Wheeler Aligner (BWA) tool was used to perform the sequence alignment to the reference genome (hg19, UCSC Genome Browser) [37]. Variant calling was performed with Genome Analysis Toolkit (GATK) Unified Genotyper [45]. Filtering of the data occurred using the following criteria: no occurrence or a frequency ≤ 0.5% of the variants in public exome variant repositories (Exome Aggregation Consortium, 1000 Genomes Project, Exome Variant Server, in‐house data); variants with impact on the encoded protein (missense, nonsense, frame shift, in-frame insertions/deletions and splice site variants); read depth ≥ 7; minimal heterozygous call percentage ≥ 20%, minimal homozygous call percentage ≥ 66%. For possible pathogenic variants, Sanger sequencing was used for validation and segregation analysis in all available family members.

#### Family D

Informed consent for diagnostic and research studies was obtained as approved by the central institutional review board at the NIH National Human Genome Research Institute for the Undiagnosed Diseases Network (UDN). Trio whole genome sequencing was performed through the UDN on genomic DNA of D:II:1, D:I:1 and D:I:2 (Fig. 1) using previously described methods [63]. Sequencing was performed using the Illumina HiSeq X sequencing platform. After sequencing, reads were generated using Illumina’s bcl2fastq and data were aligned to the human reference GRCh37. Sequence variants were loaded into a custom software analysis application called Codicem for interpretation. Genome sequencing was initially non-diagnostic. However, UDN-developed machine-learning based copy number variant bioinformatic pipelines identified an apparently *de novo* heterozygous 500 base pair deletion encompassing exon 9 of *HNRNPA1* (NM_002136.4: c.907+15_∗5-68del) [10,23]. The deletion was orthogonally confirmed via a quantitative-PCR assay. Transcriptome sequencing (RNA-seq) was performed on D:II:1 cultured fibroblasts to assess the effect of the deletion on the *HNRNPA1* mRNA transcript using previously described methods [14]. RNA-seq revealed skipping of exon 9 (https://varsome.com/transcript/hg19/NM_002136.4) predicted to result in a truncated protein [p.G304Nfs∗3] (Fig. 2, A1). In addition, RNA-seq did not show a decrease in *HNRNPA1* mRNA expression levels when compared to controls, indicating the aberrant transcript escapes from nonsense mediated decay (NMD) (Fig. A1).

**Fig. 2.**
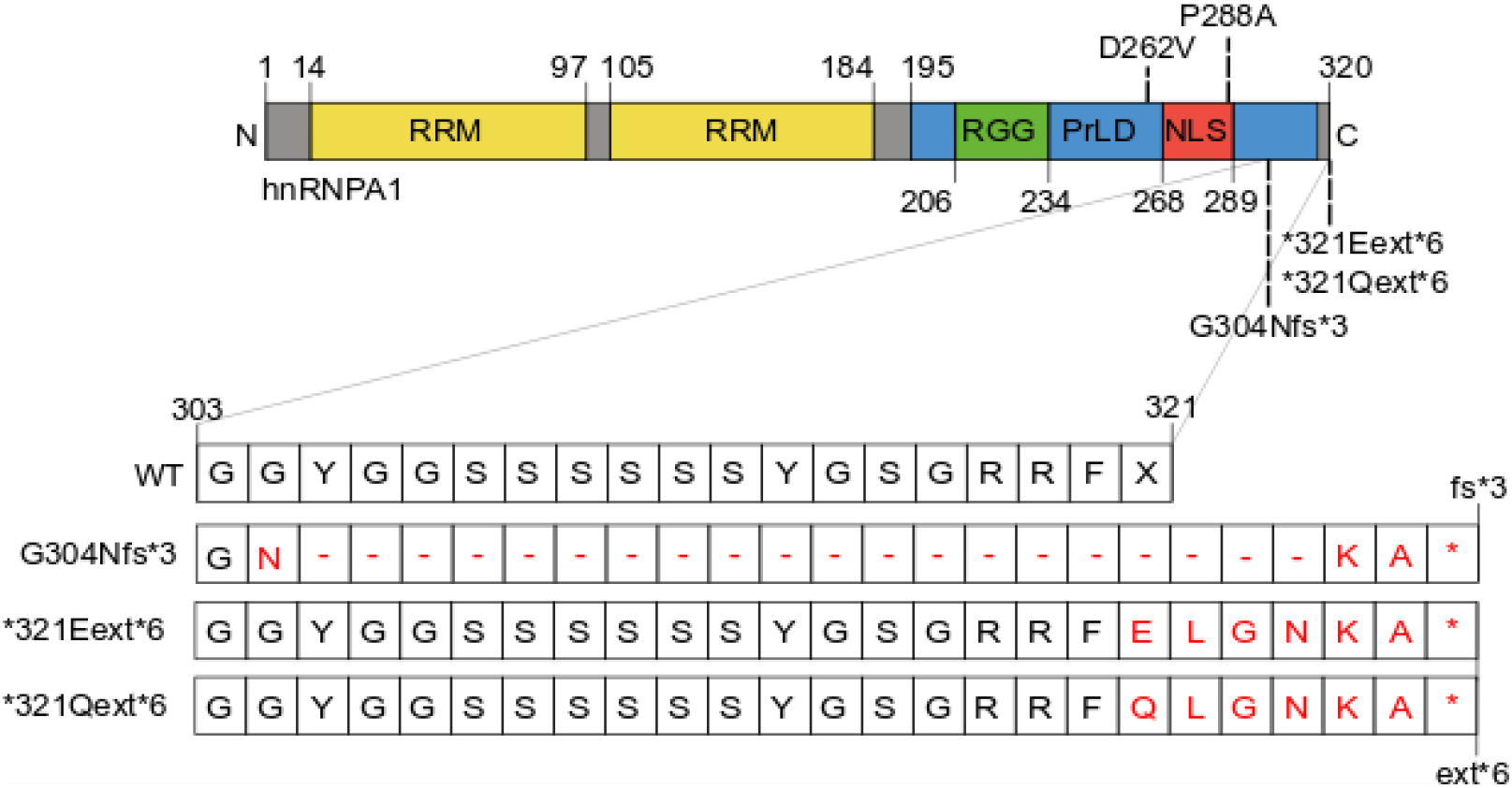
Schematic overview of hnRNPA1 protein and the effect of the mutations. hnRNPA1 contains two N-terminal RNA-recognition motifs (RRMs) and a C-terminal prion-like domain (PrLD), containing an RGG motif and a PY-NLS that enables nuclear import. Mutations studied in this paper are depicted at the appropriate location. The frameshift and extension G304Nfs∗3, ∗321Eext∗6, and ∗321Qext∗6 mutations’ effects at the protein level are shown in the zoom-in of amino acid 303-321. Single letter annotation for amino acid residues is used.

#### Family E

Whole-exome sequencing was performed on high molecular weight genomic DNA of E:I:2 and E:II:1 (Fig. 1). Exome enrichment was performed using SureSelect Clinical Research Exome V2 (Agilent). Subsequent sequencing was performed using a NovaSeq 6000 Illumina platform. The Burrows‐Wheeler Aligner (BWA) tool was used to perform the sequence alignment to the reference genome (hg19, UCSC Genome Browser) [37]. Primary data analysis is performed using Illumina DRAGEN Bio-IT Platform v.2.03. Secondary and tertiary data analysis is performed using PerkinElmer’s internal ODIN v.1.01 software for SNVs and Biodiscovery’s NxClinical v.4.3 or Illumina DRAGEN Bio-IT Platform v.2.03 for CNV and absence of heterozygosity (AOH). Variants are called at a minimum coverage of 8X and an alternate allele frequency of 20% or higher. Single nucleotide variants (SNVs) meeting internal quality assessment guidelines are confirmed by Sanger sequence analysis. All differences from the reference sequences (sequence variants) were assigned to one of five interpretation categories (Pathogenic, Likely Pathogenic, Variant of Uncertain Significance, Likely Benign and Benign) per ACMG Guidelines [57].

#### Family F

Whole-exome sequencing (WES) was performed on high molecular weight genomic DNA sample of F:I:1 (Fig. 1). The Twist Human Core Exome kit supplemented with additional probes for human RefSeq transcripts was used for exome enrichment (Twist Bioscience), with subsequent library sequencing on a NextSeq 500/550 Illumina platform. The Burrows‐Wheeler Aligner (BWA) tool was used to perform the sequence alignment to the reference genome (hg19, UCSC Genome Browser) [37]. Variant filtering was performed using VariantDB [67] using the following criteria: no occurrence or a frequency ≤ 2% of the variants in public exome/genome variant repositories (Genome Aggregation Database, gnomAD)); variants with impact on the encoded protein (missense, nonsense, frameshift, inframe insertions/deletions, splice site, start and stop loss variants); read depth ≥ 5; minimal allelic ratio ≥ 15%. Analysis of variants was limited to genes listed in a predefined myopathy gene panel (supplementary information) supplemented with genome-wide analysis based on HPO term associations (MOON software, Diploid/Invitae). Sanger sequencing was used for confirmation of WES identified variants that did not meet following criteria: read depth ≥ 30 or allelic ratio between 40 and 60% for heterozygous variants.

### *HNRNPA1* variant screening

A Multiplex Amplification of Specific Targets for Resequencing (MASTR™, Agilent) assay was designed for the mutation screening of all coding regions and exon-intron boundaries of the *HNRNPA1* gene in an additional 547 patients. Both the multiplex PCRs and following barcoding step were carried out on a Veriti AB machine (Life Technologies). Libraries were sequenced on a MiSeq Platform (Illumina) using the v3 chemistry with read lengths of 2×300 bp. For the read alignment, variant calling, annotation and filtering GATK, SAMtools and GenomeComb were used as previously described. For possible pathogenic *HNRNPA1* variants, Sanger sequencing was used for validation and segregation analysis in all available family members.

### Plasmid generation

mRNA for a control individual, index patient family A, and index patient family C was extracted from lymphoblast cell lines by RNeasy^®^ Mini Kit (Qiagen). cDNA was generated by reverse transcription using the SuperScript^®^ III First-Strand Synthesis System (ThermoFisher Scientific). cDNA was used for the PCR amplification of *HNRNPA1* (NM_002136.3) with primers containing sequences suitable for subsequent Gateway^®^ Cloning Technology. pDONR221 was used as entry vector. The P288A variant from family B was introduced into the wild-type (WT) *HNRNPA1* entry clone by site-directed mutagenesis [71]. The G304Nfs∗3 mutant from family A and D and the ∗321Eext∗6 variant from family C were cloned by direct amplification of the patient cDNA. The mRNA for the variant from family D is the same as the variant as for family A, and as such, the same plasmid was used. The entry clones were subcloned into a pLenti backbone with N-terminal V5- or EmGFP-tag for stable transduction and puromycin selection marker. Templates were checked by Sanger sequencing.

Yeast plasmids with each hnRNPA1 variant were generated by cloning the protein into the pAG416GAL-GFP-ccdB vector backbone using Gateway reactions (ThermoFisher). Mutations were generated via QuikChange Site-Directed Mutagenesis Kit (Agilent) according to the manufacturer’s instructions. Mutations were verified by DNA sequencing. Plasmids for bacterial protein expression were generated by cloning into a pDUET-GST vector backbone and mutations were made via QuikChange Site-Directed Mutagenesis Kit (Agilent) according to the manufacturer’s instructions. Mutations were verified by DNA sequencing.

### Western blot

Henrietta Lacks’ (HeLa) cells were incubated at 37 °C in a humidified atmosphere with 5% CO_2_. Cells were cultured in Minimum Essential Medium (MEM), with L-glutamine and Earle’s salts (ThermoFisher Scientific) with 10% heat-inactivated fetal bovine serum, 1% L-glutamine, 1% penicillin-streptomycin. Transient transfection of HeLa cells with the wild-type and mutant HNRNPA1 lentiviral plasmids was performed using polyethylenimine (PEI). Transfected HeLa cells were cultured for 24 h before collection.

After transfection, HeLa cells (4 x 10^6^) were pelleted and lysed with RIPA lysis buffer (150 mM NaCl, 0.5% sodium deoxycholate, 0,1% sodium dodecyl sulphate [SDS], 1% NP-40, 50 mM Tris-HCl pH 7.4) supplemented with protease inhibitors (Roche Diagnostics-Applied Science). 12.5µg of total protein was run on NuPAGE^®^ Novex^®^ 4–12% Bis-Tris Protein Gel (ThermoFisher Scientific). Primary antibody mouse anti-V5 (1:5000; Life Technologies R960-25) and secondary goat anti-mouse IgG_2a_horseradish peroxidase (Southern Biotech; diluted 1:10000) were used and visualization was performed with enhanced chemiluminescence detection (PierceTM ECL Plus Western Blotting Substrate, ThermoFisher Scientific). Equal loading was assessed by primary rabbit antibody α-tubulin (Abcam Rabbit polyclonal IgG (ab4074) diluted 1:5000) with secondary goat anti-rabbit IgG horseradish peroxidase (Southern Biotech; 1:10000).

Yeast cells transformed with the appropriate plasmid were grown in galactose-containing media to induce protein expression for 8 h. Cultures were normalized to an OD_600_= 0.6 and 6 mL of cells were harvested. Yeast lysates were extracted by incubation with 0.1 M NaOH at room temperature for 5 min. Lysates were mixed with SDS sample buffer, boiled for 5 min, and subjected to Tris-HCl SDS-PAGE (4-20% gradient, Bio-Rad) followed by transfer to a PVDF membrane (Millipore). Membranes were blocked in Odyssey blocking buffer (LI-COR) for 1 h at room temperature. Primary antibody incubations were performed at 4 °C overnight. After washing with PBS with 1% Tween (PBST), membranes were incubated with fluorescently labeled secondary antibodies at room temperature for 1 h, followed by washing with PBST. Proteins were detected using an Odyssey Fc Dual-Mode Imaging system. Antibodies used: rabbit polyclonal anti-GFP (Sigma-Aldrich), mouse monoclonal anti-PGK1 (ThermoFisher), fluorescently labeled anti-mouse and anti-rabbit secondary antibodies (Li-Cor).

### Yeast transformation and spotting assays

All experiments were performed using *S. cerevisiae* strain BY4741 (MATa, his3Δ1, leu2Δ0, met15Δ0, uraΔ30). Yeast procedures were performed according to standard protocols [18]. The PEG/lithium acetate method were used to transform yeast with plasmid DNA [24]. For spotting assays, yeast cells were grown overnight at 30 °C in liquid media containing raffinose (SRaf/-Ura) until they reached log or mid-log phase. Cultures were then normalized for OD_600_, serially diluted, and spotted onto synthetic solid media containing glucose, sucrose/galactose (1:1), or galactose lacking uracil and were grown at 30 °C for 2-3 days. Yeast experiments were performed in triplicate.

### Protein purification

hnRNPA1 proteins were purified as described [65,26]. Briefly, WT and mutant hnRNPA1 proteins were expressed and purified from *E. coli* as GST-tagged proteins. hnRNPA1 with N-terminal GST tag were over-expressed into E. coli BL21(DE3)-RIL (Invitrogen). Bacteria were grown at 37 °C until reaching an OD_600_ of ∼0.6 and expression was induced by addition of 1 mM isopropyl 1-thio-β-D-galactopyranoside (IPTG) for 15-18 h at 15 °C. Protein was purified over a glutathione-Sepharose column (GE) according to manufacturer’s instructions. GST-hnRNPA1 was eluted from the glutathione Sepharose with 40 mM HEPES-NaOH, pH 7.4, 150 mM potassium chloride, 5% glycerol, and 20 mM reduced glutathione. After purification, proteins were concentrated to ≥10 μM using Amicon Ultra-4 centrifugal filter units (10 kDa molecular weight cutoff; Millipore). Protein was centrifuged at 16,100 g for 10 min at 4 °C to remove any aggregated material before use. After centrifugation, the protein concentration in the supernatant was determined by Bradford assay (Bio-Rad) and these proteins were used for fibrillization or LLPS assays.

### *In vitro* fibrillization assays

hnRNPA1 (5 µM) fibrillization (100 µl reaction) was initiated by addition of 1 μl of 2 mg/ml TEV protease in A1 assembly buffer (40 mM HEPES-NaOH, pH 7.4, 150 mM KCl, 5% glycerol, 1 mM DTT, and 20 mM glutathione). The hnRNPA1 fibrillization reactions were incubated at 25 °C for 24 h with agitation at 1,200 rpm in an Eppendorf Thermomixer at which time fibrillization was complete with ∼100% of the hnRNPA1 in the aggregated state. For sedimentation analysis, at indicated time points, fibrillization reactions were centrifuged at 16,100g for 10 min at 24 °C. Supernatant and pellet fractions were then resolved by SDS-PAGE and stained with Coomassie Brilliant Blue, and the relative amount in each fraction was determined by densitometry in ImageJ (NIH). All fibrillization assays were performed in triplicate. For electron microscopy, fibrillization reactions (10 μl) were adsorbed onto glow-discharged 300-mesh Formvar/carbon coated copper grids (Electron Microscopy Sciences) and stained with 2% (w/v) aqueous uranyl acetate. Excess liquid was removed, and grids were allowed to air dry. Samples were viewed by a JEOL 1010 transmission electron microscope.

### *In vitro* LLPS assays

hnRNPA1 liquid droplets were formed by incubating hnRNPA1 in the absence of TEV protease at the indicated concentration in LLPS buffer (40 mM HEPES-NaOH pH 7.4, 150 mM NaCl, 5% glycerol, 1 mM DTT, and 20 mM glutathione, 10% Dextran) at 25 °C. Protein samples were spotted onto a coverslip and imaged using a Leica DMI6000 by differential interference contrast (DIC) microscopy.

### Immunofluorescence

HeLa cells were seeded on 8-well glass slides (Millipore). Cells were transfected 24 h after seeding using FuGENE 6 (Promega) with either EGFP-tagged or EYFP-tagged hnRNPA1 constructs. 24 h post transfection, cells were stressed with 500 μM sodium arsenite (Sigma-Aldrich) for 30 min. Cells were then fixed with 4% paraformaldehyde (Electron Microscopy Sciences), permeabilized with 0.5% Triton X-100, and blocked in 5% bovine serum albumin (BSA). Primary antibodies used were against G3BP1 (611127; BD Biosciences) and eIF3η (sc-16377; Santa Cruz). For visualization, the appropriate host-specific Alexa Fluor 555 or 647 (Molecular Probes) secondary was used. Slides were mounted using Prolong Gold Antifade Reagent with DAPI (Life Technologies). Images were captured using a Leica TCS SP8 STED 3X confocal microscope (Leica Biosystems) with a 63X objective. Fluorescent images were quantified by automated puncta analysis using CellProfiler software (Broad Institute). Cells were segmented using DAPI and G3BP1 channels and granules were identified using both G3BP1 and eIF3η channels. Integrated intensities of nucleus, cytoplasm, and granules were measured.

Yeast cells that had been transformed with the indicated GFP-tagged hnRNPA1 protein or vector were grown for 8 h in galactose-containing media, pelleted, and imaged using a Leica-DMIRBE microscope before being processed using ImageJ (NIH).

### Live-cell time-lapse imaging

tdTomato-G3BP1 knock-in U2OS cells were seeded in glass-bottomed Poly-D-lysine-coated 35-mm dishes (MatTek Corporation). Cells were transfected 24 h after seeding using FuGENE 6 (Promega) with EGFP-tagged or EYFP-tagged hnRNPA1 constructs. 48 h post transfection, the dish was transferred to an Opterra II swept-field confocal microscope (Bruker) system with a stage top incubator with 5% CO2 and 60X objective with an objective heater (Bioptechs), both preheated to 37 °C. The system was left to equilibrate for 5-10 min before imaging was initiated.

For time-lapse imaging, using PrairieView software with perfect focus engaged, multipoint images were taken every 30 s with both 488 and 560 nm lasers. 2 min into imaging, the objective temperature was raised to 43 °C to begin heat shock. 30 min or 60 min later, the temperature was lowered back to 37 °C to alleviate stress and cells were imaged until granules disappeared or after 2-3 h passed. Cells with equivalent expression levels of hnRNPA1 were selected for further analysis.

### Live-cell analysis

Live-cell imaging analysis was performed manually. Only hnRNPA1-positive viable cells that did not have td-Tomato G3BP1-positive granules prior to the 2-min mark in the video were evaluated. Cells were considered granule-negative at the beginning of the video and were considered granule-positive at the frame in which distinct cytoplasmic granules were visible. The frame at which cytoplasmic granules were no longer visible was defined as the time point at which cells were again SG-negative. Images were analyzed using ImageJ (NIH). The percentage of cells that were considered granule-positive was determined and graphed at each timepoint. Two-way ANOVA with Dunnett’s multiple comparisons test was performed using GraphPad Prism 6. P < 0.05 was considered significant.

## RESULTS

### Identification of *HNRNPA1* mutations in six separate families

Multiple reports have highlighted that mutations to the *HNRNPA1* gene can result in a range of neurodegenerative disorders [6,15,16,25,32,35,42,51]. For example, in a patient from family A with motor neuropathy, a heterozygous *de novo* splice site variation (c.908-2A>G) upstream of the last exon of *HNRNPA1* was identified (Fig. 1). cDNA analysis in this patient by PCR and Sanger sequencing revealed skipping of the last exon without undergoing nonsense-mediated mRNA decay, resulting in a truncated protein [p.G304Nfs∗3] (Figs. 1, 2, A1). To systematically screen for additional hnRNPA1 mutations in patients with other similar diseases, we developed a Multiplex Amplification of Specific Targets for Resequencing (MASTR™, Agilent) assay. Employing this assay, we surveyed a large cohort of 547 patients with dHMN, CMT2, CMTint, SMA, ALS, HSP, and myopathy. From this screen, we identified a previously published missense variation (c.862C>G [p.P288A]) in the index patient of family B, which was previously shown to be causal in ALS (Fig. 1) [51]. Consequently, we performed whole-exome sequencing (WES) for this family, but this did not reveal segregating mutations in other genes known for motor neuron diseases or related disorders. Therefore, the P288A mutation is the most likely cause of disease in family B.

Independently, four additional *HNRNPA1* mutations were identified in families C-F (Fig. 1). Whole-genome sequencing (WGS) in family C revealed a single-nucleotide variant in the *HNRNPA1* stop-codon (c.961T>G [p.∗321Eext∗6]), resulting in a stop-loss mutation without undergoing nonsense-mediated mRNA decay, adding an extra six amino acids to the protein before creating an alternative stop-codon (Fig. 2). Both the stop-loss mutation and the splice variant from family A terminate on the same TAG sequence present in the 3’UTR (Fig. 2). In family D, a 500 bp deletion in *HNRNPA1* was identified through trio WGS. Interestingly, this variant results in the same mRNA alteration as the variant in family A, consequently leading to the same mutant protein [p.G304Nfs∗3]. For family E, WES in individual E:II:2 identified the known *HNRNPA1* (c.785A>T; p.D262V) missense mutation, which was previously reported in a family with autosomal dominant myopathy and Paget’s disease of bone (PDB) [32,16]. Late in the study, we identified a nearly identical mutation to ∗321Eext∗6 that causes similar phenotype in the isolated index of family F (p.∗321Qext∗6) (Figs. 1, 2; Table 1). While functional studies of this ∗321Qext∗6 variant have not been performed, it is likely that this variant behaves similar to the ∗321Eext∗6 mutation.

### *HNRNPA1* mutations lead to a wide variety of different phenotypes

#### Family A - (p.G304Nfs∗3)

In his teens, the isolated patient (A: II:1) presented their first symptoms with wasting of small hand muscles. A year after onset, sural nerve biopsy showed very mild axonal neuropathy with secondary segmental demyelination. Electromyography (EMG) in median, ulnar, and peroneal nerves showed decreased amplitudes of compound muscle action potentials (CMAPs) with nerve conduction velocities (NCVs) in the normal range, while no abnormalities were seen in sensory fibers. In his 20’s, he displayed right-sided thoracic scoliosis, mild *pes cavus*, wasting of the forearms and calves (1/3 lower part), and brisk deep tendon reflexes in the upper and lower limbs with intact sensory modalities. In his 40’s, he had severe wasting of the distal muscles of the upper and lower limbs. Despite his very clumsy gait with foot drop, he is still ambulant. Overall, his phenotype corresponds to an axonal motor predominant neuropathy both in the upper and lower limbs with wasting of the first dorsal interosseous muscle and atrophy of the lower parts of the forearms. Additionally, mild pyramidal signs are present with very brisk knee reflexes and brisk ankle reflexes. From a clinical point of view, this patient’s symptoms fit within the spectrum known for dHMN.

#### Family B - (p.P288A)

Patient B:III:1 is a woman in her 40’s, who developed painless, proximal right arm weakness and wasting, without sensory disturbances in her 20’s. Two higher generation relatives presented the same phenotype. Both of whom died from respiratory failure around the age of 40. Her diagnosis of a very slowly progressive type of motor neuron disease was made four years after symptom onset, based on the clinical features together with supportive neurophysiological findings: upper and lower motor neuron symptoms observed at the clinical examination and a chronic neurogenic pattern limited to the upper limb and tongue muscles recorded via EMG. Over time, she developed progressive weakness and wasting of the left arm, respiratory insufficiency, and lower limb spasticity. Five years after symptom onset, she developed bulbar symptoms with dysarthria, rhinolalia and chewing difficulties, and eventually dysphagia for both liquids and solids. She required percutaneous endoscopic gastrostomy three years later. After about 20 years of disease progression, she is still walking without aid. Three relatives B:IV:1-3 are presently unaffected all at around 20 years of age. At around 40 years of age, individual B:III:2 presented with progressive bulbar symptoms including dysarthria, tongue atrophy, and swallowing problems. She also presented upper motor neuron signs, and right hand weakness with amyotrophy.

#### Family C - (p.∗321Eext∗6)

Patient C:II:2 is showed her first symptoms before the age of 10. She presented with difficulties with handgrip while writing, mild facial weakness, and general fatigue in her teens. Muscle weakness and atrophy was gradually progressive. She has been wheelchair bound since her early 30’s. In her early 40’s, muscle strength in both upper and lower limbs had become severely reduced, and reflexes were absent in all limbs. She also had a very severe facial weakness and subsequent dysarthria. An M. quadriceps biopsy was performed, showing non-specific myopathic changes with some angular fibers, but no type grouping. Nerve conduction studies (NCS) were normal apart from small CMAPs due to severe distal atrophy. Needle EMG was compatible with a myogenic process. The patient developed a mild dissociated vital hyposensitivity distally in the legs, but no abnormalities were noted on spinal MRI. A clinical diagnosis of facio-scapulo-humeral muscular dystrophy (FSHD) was made, but could not be confirmed genetically.

Individual C:III:1 presented his first symptoms with a right-sided foot drop before the age of 10 as well. In his teens he is still ambulant and can still walk on his toes. However, heel-walking is impossible. He shows slight wasting of the left thenar and reduced muscle strength in the lower limbs. His reflexes are normal in all limbs. NCS were normal and needle EMG revealed myopathic features. A needle biopsy of the M. quadriceps was performed at age 8, which showed mild and non-pathognomonic myopathic features. Overall, the phenotype in this family is compatible with a myopathy with distal and facial onset and severe proximal and bulbar weakness upon progression. The distal weakness, atrophy and subsequent small CMAPs suggested an overlap with the dHMN disease spectrum but the myopathic features initially dominated the phenotype.

#### Family D - (p.G304Nfs∗3)

Individual D:II:1 is a is a woman in her early 30’s, who developed right hand weakness, pain when writing, and atrophy since her early 20’s. She was thought to have multifocal motor neuropathy based on her EMG, but was treated with a trial of intravenous immunoglobin (IVIG) without response. Her muscle biopsy revealed vacuolar myopathy, notable for its myofibrillar disorganization, moth-eaten fibers, rimmed vacuoles and an absence of inflammation. Repeat EMG around age 30 showed irritative distal myopathy. Her symptoms have been gradually progressive: in her late 20’s, she noted right ankle weakness, which is now bilateral. Her exam is notable for mild right facial weakness and her distal weakness is markedly asymmetric: right finger and wrist extension was 1/5 on the R, 4 to 4+ on the left, and right ankle dorsiflexion 0/5, L is 4-/5. Deep finger flexor strength is spared. Reflexes were initially preserved in the lower extremities (+2 at presentation) but have diminished over time. There is no spasticity nor upper motor neuron signs. She walks unassisted, has scapular winging, and has reported mild swallowing difficulties with normal speech. Her spirometry shows a restrictive pattern with predicted forced vital capacity (FVC) of 76%, and a sleep study showed sleep-disordered breathing. Her creatine kinase (CK) has been mildly elevated (CK range: 192-315 U/L). Cardiac MRI, electrocardiogram and fourteen-day cardiac rhythm monitor were all normal.

#### Family E - (p.D262V)

Individual E:II:2 is a male, who developed asymmetric weakness of the right foot in his 30’s that worsened over time to involve bilateral proximal leg weakness. He initially noticed difficulties with snow skiing. He started to have frequent tripping and falls, leading to multiple falls per week. Over the course of 10 years there was slow progression of bilateral leg weakness that now requires use of a cane for ambulation. He also developed mild difficulty with dressing, hygiene and lifting objects mainly due to leg and balance problems. Currently, in his late 40’s the patient uses his arms to stand from sitting position, and requires use of hand rails when going up the stairs. He reports no weakness of facial, neck, arm or trunk muscles; numbness or tingling; dysphagia; difficulty with hand-writing, handling utensils or opening doors; shortness of breath; bowel/bladder dysfunction; diplopia; or muscle pain. NCS abnormalities were limited to low amplitude or the right fibular compound muscle action potentials (CMAPs), whereas needle EMG revealed chronic myopathic changes with muscle membrane irritability in the proximal and distal muscles of the leg. A muscle biopsy at age 45 showed chronic myopathy with rimmed vacuoles.

The patient’s family history is significant for debilitating muscle disease as his relative (E:I:2) was diagnosed with Paget’s disease of bone (PDB), when was she found to have elevated alkaline phosphatase on her annual checkup in her late 40’s. Within 3 years she developed left leg weakness that gradually progressed to involve both legs and currently, she is wheelchair bound. She has normal arm strength and normal cranial nerve function and a muscle biopsy was consistent with myopathy with inclusion bodies. The index patient has six unaffected same generation relatives in an age range of 30-50 years. The overall phenotype in this family is consistent with inclusion body myositis.

#### Family F - (p.∗321Qext∗6)

Patient F:I:1 is a male, who presented his first symptoms in his early teens. He presented with muscle atrophy in the hands, with subsequent progressive wasting of the lower limbs starting distally before also involving proximal muscles. At the last examination in his 60’s, this patient was wheelchair-bound with profound generalized weakness resulting in *de facto* quadriplegia. Reflexes are lost in all extremities. In addition to contractures in the lower limbs and bilateral *pes equinovarus*, the patient also displayed marked facial weakness. Respiration is severely limited requiring cough assist and non-invasive home ventilation (BiPAP). The patient also makes use of a percutaneous endoscopic gastrostomy (PEG). A biopsy of the M. tibialis anterior in his teens showed myogenic alterations consisting of marked diameter variation, selective type 1 fiber atrophy, increased centralized nuclei and the presence of rimmed vacuoles. A biopsy of the N. fibularis superficialis was strictly normal. Nerve conduction studies and needle EMG did not show neurogenic changes and were otherwise in keeping with a distally predominant myopathy. The patient has since passed away shortly after the last examination. The family history for this patient is negative for neuromuscular disorders, making this an isolated case. The overall clinical presentation fits with a progressive myopathy with onset in the distal limbs. A clinical diagnosis of distal myopathy of Welander type or Nonaka type was suggested. UDP-N-acetylglucosamine-2-epimerase/N-acetylmannosamine kinase (*GNE*) mutation analysis remained negative.

### Comparable in vitro transcription of hnRNPA1 mutants

To verify that the alternatively terminating variants were not subjected to nonsense-mediated decay (NMD), mRNA sequences for the G304Nfs∗3 (Family A) and ∗321Eext∗6 (Family C) variants were checked by PCR amplification of exon 8 into the 3’UTR on cDNA from patient lymphoblasts. Gel electrophoresis and di-deoxy sequencing confirmed the skipping of exon 11 for the splice variant (Fig. A1a). We found that the mRNA for both the G304Nfs∗3 and ∗321Eext∗6 variants is present, showing the absence of NMD and suggesting that these genes are transcribed and potentially translated in patients (Fig. A1, b and d). To study the translation of the mutated protein in a cellular model, hnRNPA1 plasmids were constructed for N-terminally V5 tagged wild-type cDNA, P288A, G304Nfs∗3 and ∗321Eext∗6 mutations and each protein was transiently expressed in HeLa cells. Western blotting confirmed translation of wild-type hnRNPA1 and the three different mutant proteins in this model. As expected, the G304Nfs∗3 mutation produced a shorter protein, while the ∗321Eext∗6 mutation translated to a longer protein (Fig. A1e). The P288A mutation translated into a protein of equal size to the wild-type hnRNPA1 (Fig. A1e). Thus, these patient-derived mutant mRNA transcripts can be translated into mutant hnRNPA1 protein in cells.

### Wild-type and mutant hnRNPA1 confer toxicity in yeast

For additional insight into the consequences of the patient mutations described thus far, we used a yeast model to express each hnRNPA1 variant. The hnRNPA1 proline-tyrosine nuclear localization signal (PY-NLS) is not decoded by the yeast nuclear-import machinery [64]. Thus, expression of wild-type hnRNPA1 or hnRNPA1^D262V^ in yeast leads to its cytoplasmic aggregation and toxicity, which phenocopies pathological events in MSP [32]. To assess whether the novel hnRNPA1 variants were also toxic in yeast, we performed spotting assays on glucose (no expression of hnRNPA1), sucrose/galactose (1:1; moderate expression of hnRNPA1), or galactose-containing agar plates (high expression of hnRNPA1) to assess the impact of hnRNPA1 expression on cell growth. Expression of wild-type or mutant hnRNPA1 under a galactose-inducible promoter on both the sucrose/galactose (1:1) and galactose-containing agar plates led to dramatic inhibition of growth compared to cells transformed with an empty plasmid (Fig. 3a). Western blot confirmed that hnRNPA1 variants were robustly expressed in yeast when given galactose as a carbon source (Fig. 3b). Thus, in yeast, the hnRNPA1 variants display similar toxicity to wild-type hnRNPA1 (Fig. 3a). Next, we looked at the localization and aggregation status of mutant hnRNPA1. Assessment of the subcellular localization of hnRNPA1 in yeast revealed that hnRNPA1, hnRNPA1^D262V^, hnRNPA1^P288A^, and hnRNPA1^∗321Eext∗6^ form cytoplasmic foci (Fig. 3c). By contrast, hnRNPA1^G304Nfs∗3^ displayed more diffuse localization in yeast, although cytoplasmic foci could be found in some cells (Fig. 3c). Because G304Nfs∗3 is expressed to similar levels as the other mutant protein (Fig. 3b), these findings indicate that hnRNPA1^G304Nfs∗3^ may be less aggregation-prone than hnRNPA1, hnRNPA1^D262V^, hnRNPA1^P288A^, and hnRNPA1^∗321Eext∗6^.

**Fig. 3.**
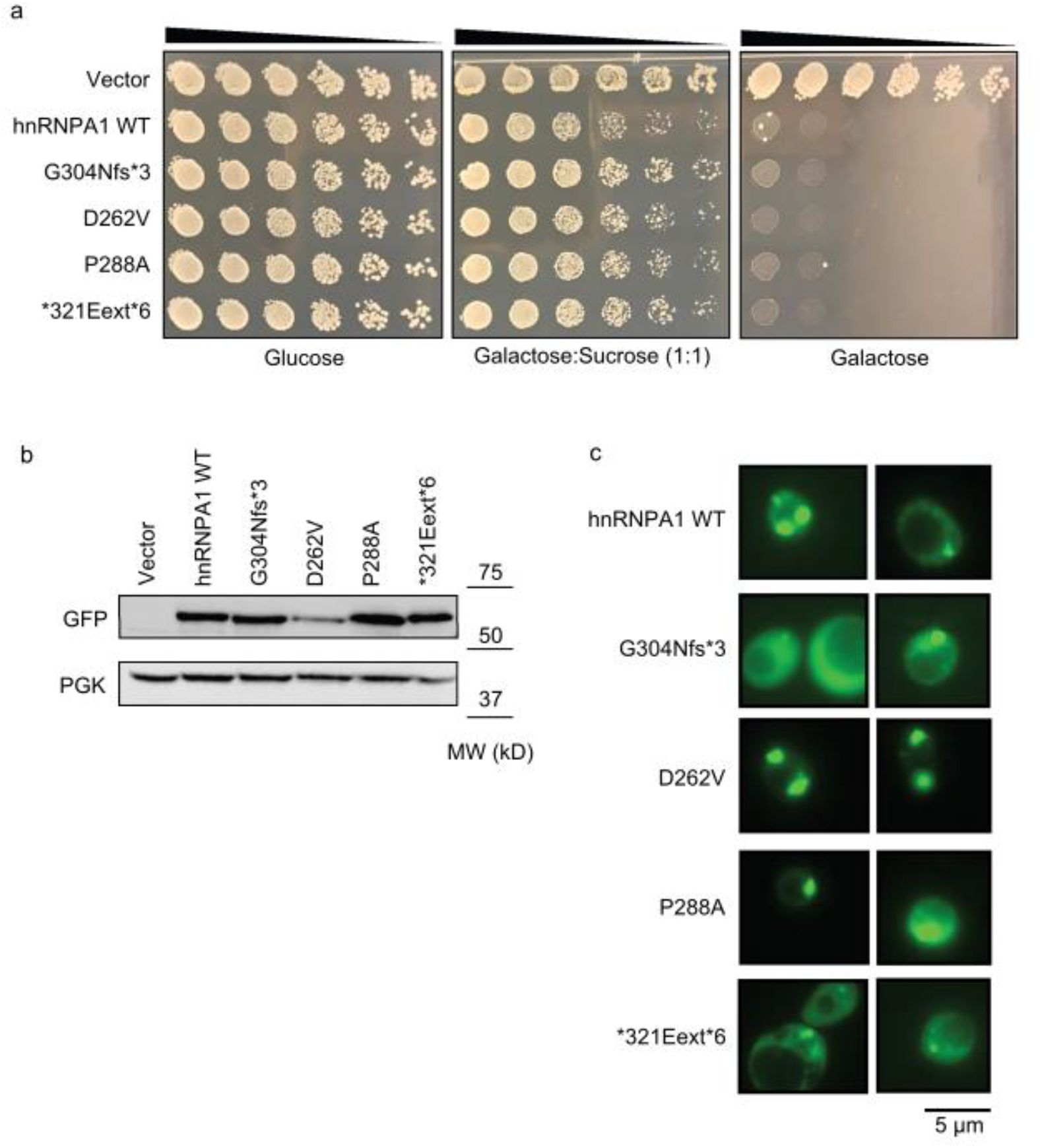
hnRNPA1 variants are toxic and aggregate in yeast. **a**. GFP-tagged hnRNPA1 variants are toxic when expressed in yeast. Yeast spotting assays compare the toxicity of different hnRNPA1 variants by plating yeast at serially diluted concentrations. hnRNPA1 variants were expressed from a galactose-inducible promoter. Thus, expression of hnRNPA1 is repressed when cells are grown in the presence of glucose (left panel), then moderately induced in the presence of sucrose:galactose (middle panel), and strongly induced in the presence of galactose (right panel). **b**. Western blot confirmed the expression of GFP-tagged hnRNPA1 variants in yeast. PGK is used as loading control. **c**. Yeast cells expressing GFP-tagged WT and mutant human hnRNPA1. hnRNPA1, hnRNPA1^D262V^, hnRNPA1^P288A^, and hnRNPA1^∗321Eext∗6^ formed cytoplasmic foci. hnRNPA1^G304Nfs∗3^ displayed more diffuse localization in yeast, although cytoplasmic foci could be found in some cells.

### Differential fibrillization and LLPS propensity of hnRNPA1 mutants

Our yeast work suggested that hnRNPA1 mutants differ in the degree to which they self-assemble. To investigate how the mutations affect the fibrillization propensity of hnRNPA1, we first examined wild-type hnRNPA1, as well as the disease-associated mutants with ZipperDB [17]. ZipperDB is a structure-based threading algorithm, which scores six-amino-acid segments for their propensity to form two self-complementary beta strands termed ‘steric zippers’ that form the spine of amyloid fibrils. Hexapeptides with Rosetta energy of lower than −23 kcal/mol are predicted to form steric zippers, with lower energy predicting higher amyloidogenicity. ZipperDB predicted that the P288A mutation introduces a new steric zipper (285-SSGAYG-290) that could increase the fibrillization propensity (Fig. 4a). By contrast, G304Nfs∗3 deletes a large stretch of steric zipper motifs present in the PrLD of hnRNPA1, which could reduce aggregation propensity by reducing the multivalency of the PrLD (Fig. 4a). Finally, the ∗321Eext∗6 mutation did not change the steric zipper landscape, with ZipperDB predicting a similar fibrillization propensity to wild-type hnRNPA1 (Fig. 4a).

**Fig. 4.**
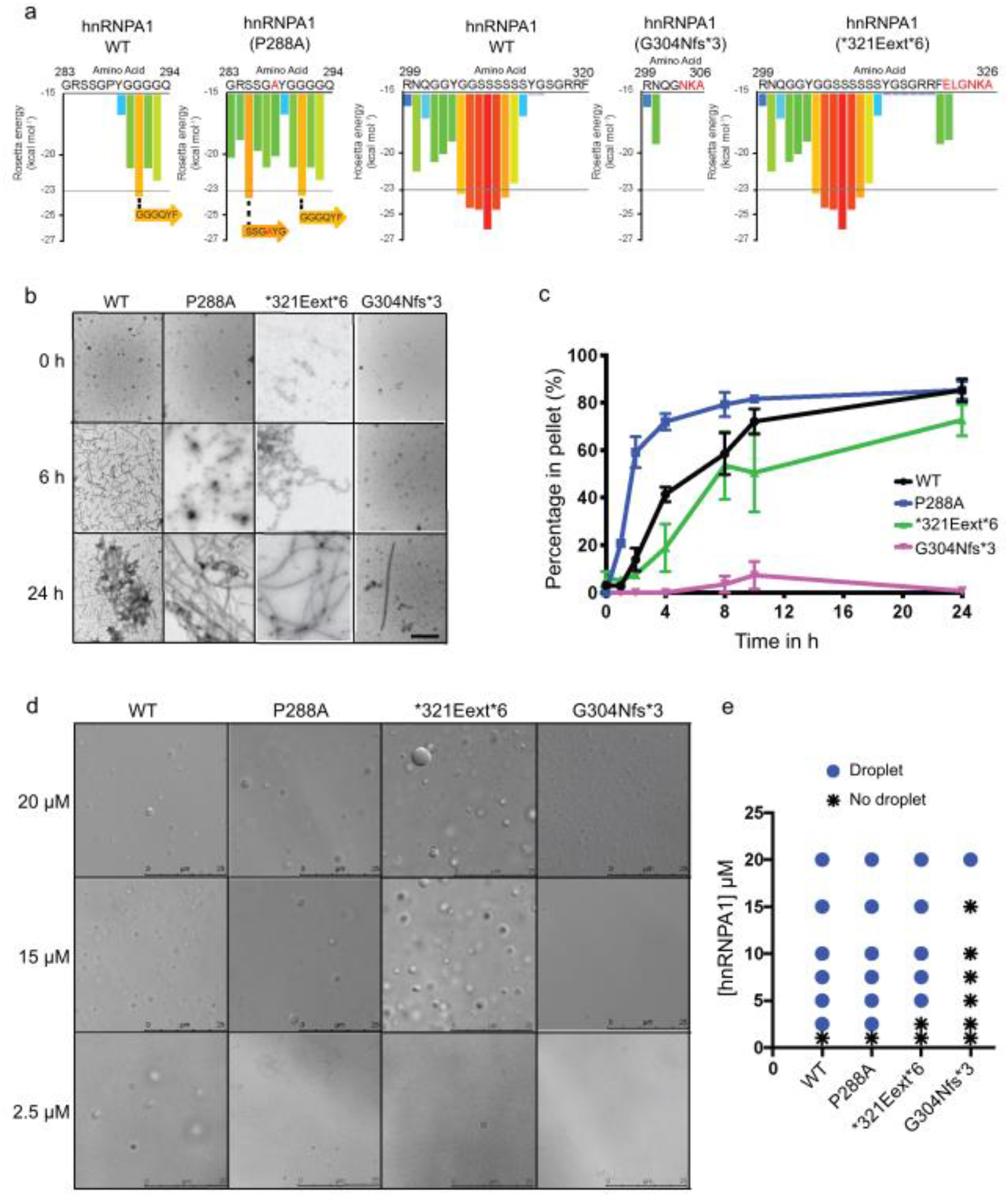
hnRNPA1 variants can exhibit altered propensity for fibrillization and liquid-liquid phase separation. **a**. ZipperDB, a structure-based algorithm, calculates the propensity of hexapeptide fragments to form steric zippers [17]. Steric zippers, which are self-complementary β-sheets that form the backbone of an amyloid fibril, are predicted to form when the Rosetta energy of a hexapeptide is below the empirically determined “high fibrillization propensity” threshold of –23 kcal/mol [17]. The P288A mutation introduces a potent steric zipper (285-SSGAYG-290) that could increase the fibrillization propensity, whereas G304Nfs∗3 deletes several potent steric zippers, which could reduce fibrillization propensity. **b, c**. GST-TEV-hnRNPA1 and disease variants (5 μM) were incubated with TEV protease in A1 assembly buffer to initiate fibrillization, and reactions were agitated at 1,200 rpm for 0-24 h at 25 °C. Fibrillization was monitored by electron microscopy (**b**). Scale bar, 0.5 µm. Alternatively, hnRNPA1 fibrillization kinetics were determined by sedimentation analysis (**c**) where the amount of hnRNPA1 in the pellet fraction was quantified. Values represent average ± SEM (n=3-6). **d**. Representative DIC microscopy images of hnRNPA1 droplets formed by different hnRNPA1 variants. hnRNPA1 and hnRNPA1^P288A^ form liquid droplets at all concentrations presented, whereas hnRNPA1^∗321Eext∗6^ does not form droplets at 2.5 μM. hnRNPA1^G304Nfs∗3^ forms droplets only at 20 μM. In addition, droplets formed by hnRNPA1^G304Nfs∗3^ are typically smaller than the droplets formed by hnRNPA1, hnRNPA1^P288A^, and hnRNPA1^∗321Eext∗6^. Scale bars, 25 µm. **e**. Phase diagram of hnRNPA1 variants showing the hnRNPA1 concentrations where LLPS occurs. hnRNPA1 and hnRNPA1^P288A^ form droplets at concentrations of 2.5 µM or higher. hnRNPA1^∗321Eext∗6^ forms droplets at concentrations of 5 µM or higher. hnRNPA1^G304Nfs∗3^ forms droplets at concentrations of 20 µM or higher.

To assess the fibrillization propensity of the different mutants in comparison to wild-type hnRNPA1, we purified GST-TEV-tagged versions of the proteins from *E. coli* [32]. Fibrillization of purified GST-TEV-hnRNPA1 was initiated by incubating with TEV protease to remove the GST tag [32]. hnRNPA1 fibrillization kinetics were then monitored by sedimentation analysis, and the final structures that formed were imaged using electron microscopy (EM) [32]. We observed that hnRNPA1^P288A^ assembled into fibrils more rapidly than hnRNPA1 (Fig. 4b, c), consistent with the prediction of ZipperDB (Fig. 4a). Thus, the proline at residue 288 likely plays a key role in disfavoring β-sheet interactions between hnRNPA1 proteins that drive fibrillization (Fig. 4a) [58,68]. Interestingly, this proline is also a critical component of the PY-NLS of hnRNPA1, which is necessary for its proper nuclear localization [61]. Therefore, in cells, the P288A mutation contributes to increased cytoplasmic mislocalization of hnRNPA1 as well as its aggregation. On the other hand, and as predicted by ZipperDB, hnRNPA1^∗321Eext∗6^ exhibited similar fibrillization kinetics as wild-type hnRNPA1 (Fig. 4b, c). Strikingly, hnRNPA1^G304Nfs∗3^ displayed greatly decelerated fibrillization kinetics, with only a few fibrils apparent after 24 h (Fig. 4b, c), again in line with the predictions made by ZipperDB (Fig. 4a) and with our observed reduction in cytoplasmic foci formation in yeast (Fig. 3c). Thus, the P288A mutation accelerates hnRNPA1 fibrillization, whereas the ∗321Eext∗6 mutation has minimal effect, and the G304Nfs∗3 reduces fibrillization.

hnRNPA1 is a component of SGs, which are liquid-like membraneless organelles that form via LLPS [32]. Similar to other PrLD-containing RBPs in SGs, purified hnRNPA1 can also form liquid droplets spontaneously upon macromolecular crowding [48]. We therefore investigated the effect of the hnRNPA1 mutations on the LLPS of hnRNPA1. In the presence of a crowding agent, WT hnRNPA1 forms liquid droplets at concentrations of 2.5 µM or greater (Fig 4d, e). Interestingly, hnRNPA1^P288A^ mutant showed similar LLPS properties to WT hnRNPA1 (Fig. 4d, e), whereas hnRNPA1^∗321Eext∗6^ had slightly reduced ability to undergo LLPS, as no droplets formed at 2.5 µM (Fig. 4d, e). These findings indicate that the features that drive fibrillization of hnRNPA1 (such as steric zippers) do not necessarily have the same effect on the formation of other self-assembled states. By contrast, hnRNPA1^G304Nfs∗3^ exhibited a marked decreased in its tendency to undergo LLPS (Fig. 4d, e). Indeed, hnRNPA1^G304Nfs∗3^ liquid droplets were only observed at 20 µM (Fig. 4d, e). Moreover, liquid droplets formed by hnRNPA1^G304Nfs∗3^ were smaller than the droplets formed by hnRNPA1, hnRNPA1^P288A^, and hnRNPA1^∗321Eext∗6^ (Fig. 4d,e). These results indicate that the G304Nfs∗3 mutation perturbs hnRNPA1 LLPS, likely by reducing the multivalency of the PrLD (Fig. 4a).

### hnRNPA1 mutants modulate stress granule dynamics

To examine the impact of disease mutations on the formation of SGs in human cells, we expressed wild-type or mutant GFP-tagged hnRNPA1 in HeLa cells and subjected these cells to environmental stressors. G304Nfs∗3 and ∗321Eext∗6 mutations did not influence the incorporation of hnRNPA1 into arsenite-induced SGs, whereas hnRNPA1^P288A^ showed significantly greater incorporation into SGs (Fig. 5a, b). This result is consistent with previous reports that mutations in the PY-NLS of hnRNPA1 interfere with its nuclear import and enhance the incorporation of hnRNPA1 into stress granules [51,46,42].

**Fig. 5.**
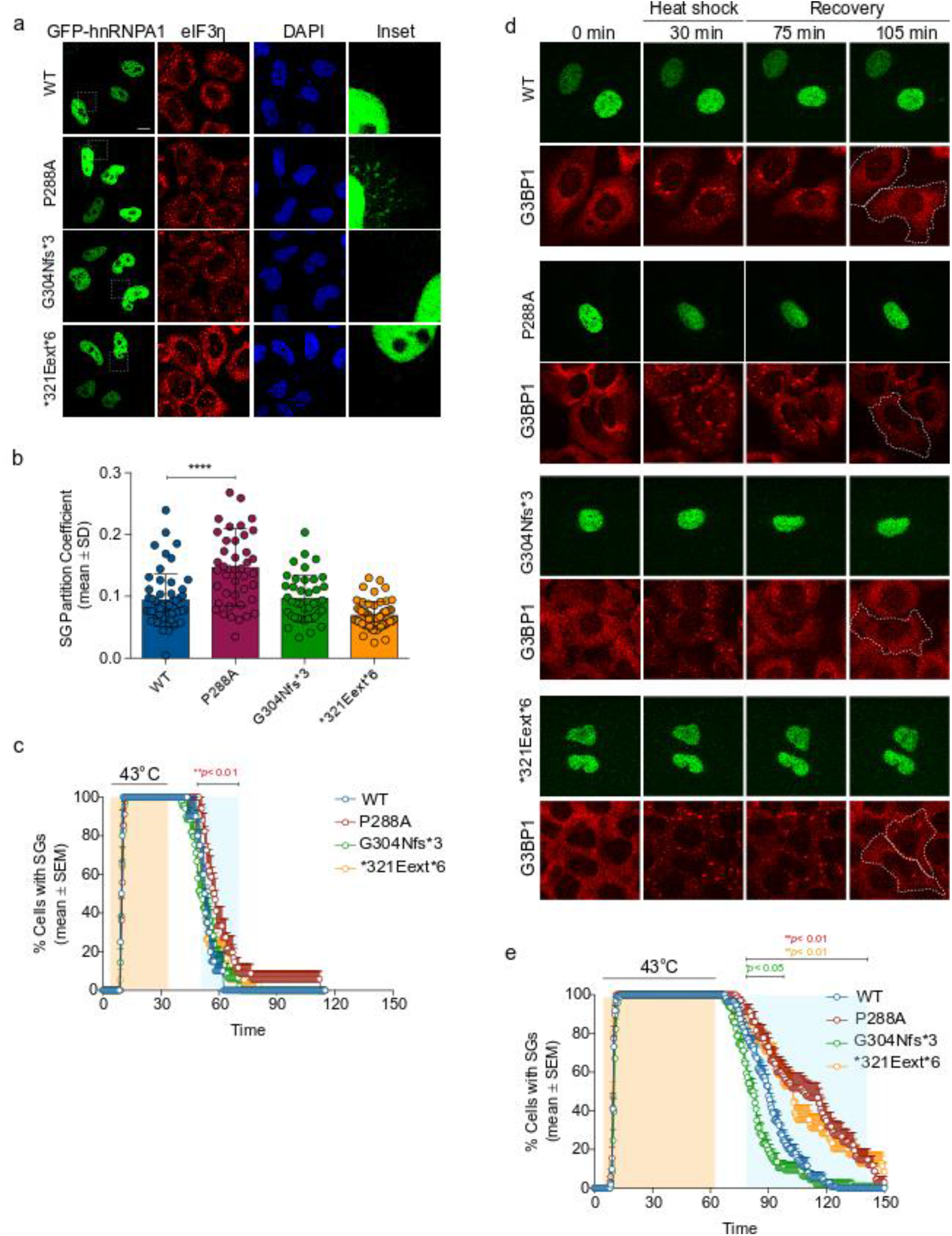
Localization and stress granule dynamics of the hnRNPA1 mutants. **a-b**. HeLa cells were transiently transfected with WT or mutant EGFP-tagged hnRNPA1 and subjected to arsenite stress (0.5 mM sodium arsenite, 30 min). Cells were fixed and stained with eIF3η (red) and DAPI (blue). Confocal images were taken for partition coefficient analysis. Error bars represent mean ± SD (n = 55, 43, 40, and 58 cells for WT, P288A, G304Nfs∗3, and ∗321Eext∗6, respectively). ∗∗∗∗p < 0.0001, ∗∗p = 0.0058 by ordinary one-way ANOVA with Dunnett’s multiple comparisons test. **c**. U2OS cells expressing tdTomato-tagged endogenous G3BP1 were transiently transfected with WT or mutant EGFP-tagged hnRNPA1 and subjected to heat shock (43°C, 30 min; orange shading) and allowed to recover at 37°C for 2 h. Line graph represents the percentage of cells with visible tdTomato-G3BP1 puncta over time. Error bars represent mean ± SEM (n = 10, 17, 23, and 18 videos for WT, P288A, G304Nfs∗3, and ∗321Eext∗6, respectively). Blue shaded area indicates time points at which P288A mutant was statistically significantly different from WT. ∗∗p < 0.01 by two-way ANOVA with Dunnett’s multiple comparisons test. **d**,**e**. U2OS cells expressing tdTomato-tagged endogenous G3BP1 were transiently transfected as in c and subjected to heat shock (43 °C, 60 min; orange shading) and allowed to recover at 37 °C for 2 h. White dotted lines delineate hnRNPA1-positive cells. Line graph in e represents the percentage of cells with visible tdTomato-G3BP1 puncta over time (n = 56, 34, 48, and 35 videos for WT, P288A, G304Nfs∗3, and ∗321Eext∗6 respectively). Blue shaded area indicates time points at which each mutant was statistically significantly different from WT. ∗p < 0.05, ∗∗p < 0.01 by two-way ANOVA with Dunnett’s multiple comparisons test. Scale bar, 10 μm.

To further assess the impact of hnRNPA1 mutations on SG dynamics, we established a live-cell assay that permitted real-time monitoring of the kinetics of SG assembly and disassembly in response to a controlled heat pulse. For this assay we used a U2OS cell line in which CRISPR/Cas9 was used to tag endogenous G3BP1 with tdTomato [69]. We then transiently transfected these cells with EGFP-tagged hnRNPA1 (wild-type or mutant forms) to permit monitoring of SG dynamics in cells expressing exogenous hnRNPA1. Although none of the hnRNPA1 mutants had a significant impact on the number or assembly rate of SGs, we did observe mutation-dependent effects on the rates of SG disassembly. When SGs were induced by a 30 min heat shock, the expression of P288A mutant slightly delayed disassembly relative to wild-type hnRNPA1 expression (Fig. 5c).

Extending the duration of the heat shock to 60 min increased the magnitude of the effects of the P288A mutant on SG disassembly and ∗321Eext∗6 mutant also showed effects on the rates of disassembly (Fig. 5d, e). While cells expressing WT hnRNPA1 or the G304Nfs∗3 mutant showed nearly complete disassembly of SGs during the 60-min recovery period (Fig. 5d, e), 50% of cells expressing either P288A or ∗321Eext∗6 mutants failed to disassemble SGs within this period (Fig. 5d, e). This delay in SG disassembly suggests that incorporation of P288A or ∗321Eext∗6 hnRNPA1 into SGs alters their dissolution. Consistent with our *in vitro* data, the G304Nfs∗3 mutant shows near complete SG disassembly and even shows faster disassembly than wild-type hnRNPA1 after prolonged heat stress.

We also evaluated how the hnRNPA1 D262V mutation, which was previously associated with MSP and identified in Family E in the current study, altered SG formation [32]. As with the other mutations tested here, the frequency of hnRNPA1 incorporation into SGs was not altered between the wild-type and D262V forms of hnRNPA1 (Fig. A2a). However, similarly to the P288A and ∗321Eext∗6 mutants, the D262V mutation did result in significantly delayed SG disassembly (Fig. A2b, c). Together, these results suggest that mutations to hnRNPA1 can have variable consequences on the rate of SG disassembly.

## DISCUSSION

Since the original report [32], only one additional *HNRNPA1* mutation was identified in a flail arm ALS family [42]. In this study, we identified and characterized both novel and previously described *HNRNPA1* mutations in atypical ALS, HMN, distal myopathy and MSP3 patients (Table 1). In six families, we identified the following mutations: stop-loss mutations ∗321Eext∗6 and ∗321Qext∗6 leading to extension of the protein; a splice site mutation and a 500bp deletion, both leading to skipping of the last exon resulting in a truncated protein G304Nfs∗3; a known missense mutation causal for ALS in an HMN family P288A; and the known MSP3 D262V variant in a myopathy family [51]. Each of these mutations are in the PrLD of hnRNPA1, suggesting that this domain has an important biological function.

Mutations in HRNNPA1 show great clinical heterogeneity. The patients in family A and B present with phenotypes of pure motor neuron disorders without sensory involvement and with pyramidal, bulbar, and upper motor neuron involvement. Striking is the overall slow progression of disease in these patients, where the patients remain ambulant even 15 to 20 years after disease onset, unlike typical ALS cases. Indeed, these patients’ symptoms are more consistent with HMN phenotypes. The patients in family C present a phenotype that is compatible with a distal onset myopathy with facial weakness with relatively faster progression, with the eldest patient having become wheelchair-bound in his 30’s. Some neurogenic features suggest an overlap with the dHMN spectrum in this family. Strikingly, the phenotype in family D is very similar to family C, rather than to family A with whom they share the same mutation. Interestingly, family F shows a myopathic phenotype with pronounced facial weakness very similar to family C, carrying the near identical mutations ∗321Eext∗6 and ∗321Qext∗6. The myopathic phenotype in family E very similar to the previously reported family carrying the D262V mutation. The identification of mutations in *HNRNPA1* in each of these families demonstrates that a broader range of phenotypes is associated with pathogenic variants in *HNRNPA1* than was previously appreciated. Despite the rarity of variants in *HNRNPA1*, our results suggest that it could be beneficial to test patients with a broader motor neuron or (distal) myopathy disease phenotype for pathogenic variants in this gene. Moreover, defining the mechanism by which hnRNPA1 mutations manifest pathologically is essential for understanding and treating diseases in which these mutations are present.

Several RNA- and DNA-binding proteins, including hnRNPA1, are implicated in neurodegenerative diseases, and many of them harbor a low-complexity PrLD [32,33,2,8,9,52,54]. Disease-associated mutations in the low-complexity PrLD of a protein can exacerbate the propensity of the entire protein to form self-seeding fibrils, resulting in accumulation of cytoplasmic aggregates and persistent SGs [39,15]. In addition to negative gain-of-function consequences to PrLD mutations, there are also loss-of-function consequences to having a mutated PrLD. In the case of hnRNPA1, the PrLD is necessary for its participation in splicing, stable RNA binding, optimal RNA annealing activity, and protein-protein interactions, and so PrLD mutations may also hinder the normal functioning of hnRNPA1 [44,59].

Here, we assessed the intrinsic fibrillization propensity for four out of the six different *HNRNPA1* mutations identified in this study (Table 2). Previously, the P288A mutation was predicted to cause accelerated fibrillization kinetics, as proline can inhibit the intrinsic fibrillization propensity of a protein by inducing a local twist (structural constraint), preventing β-sheet interactions that facilitate aggregation [58,68]. Using ZipperDB, we found that the P288A mutation results in a new potent steric zipper in the PrLD (Fig. 4a) that is predicted to cause a more rapid fibrillization. This effect is in addition to the effect of the P288A mutation on increasing cytoplasmic localization by impairing nuclear import of hnRNPA1 [42]. Indeed, the P288A mutation alters the critical proline residue of the PY-NLS of hnRNPA1, which weakens its interaction with its nuclear-import receptor, Karyopherin-β2 (Kapβ2) [20]. Thus, Kapβ2 is predicted to be less able to chaperone and disaggregate hnRNPA1^P288A^ in the cytoplasm and transport hnRNPA1^P288A^ into the nucleus [20]. Our experimental data corroborates the prediction for the P288A mutation showing increased propensity for fibrillization. Thus, enhanced intrinsic fibrillization and reduced interaction with Kapβ2 (which would ordinarily prevent fibrillization) likely combine to render P288A pathogenic.

**Table 2.**
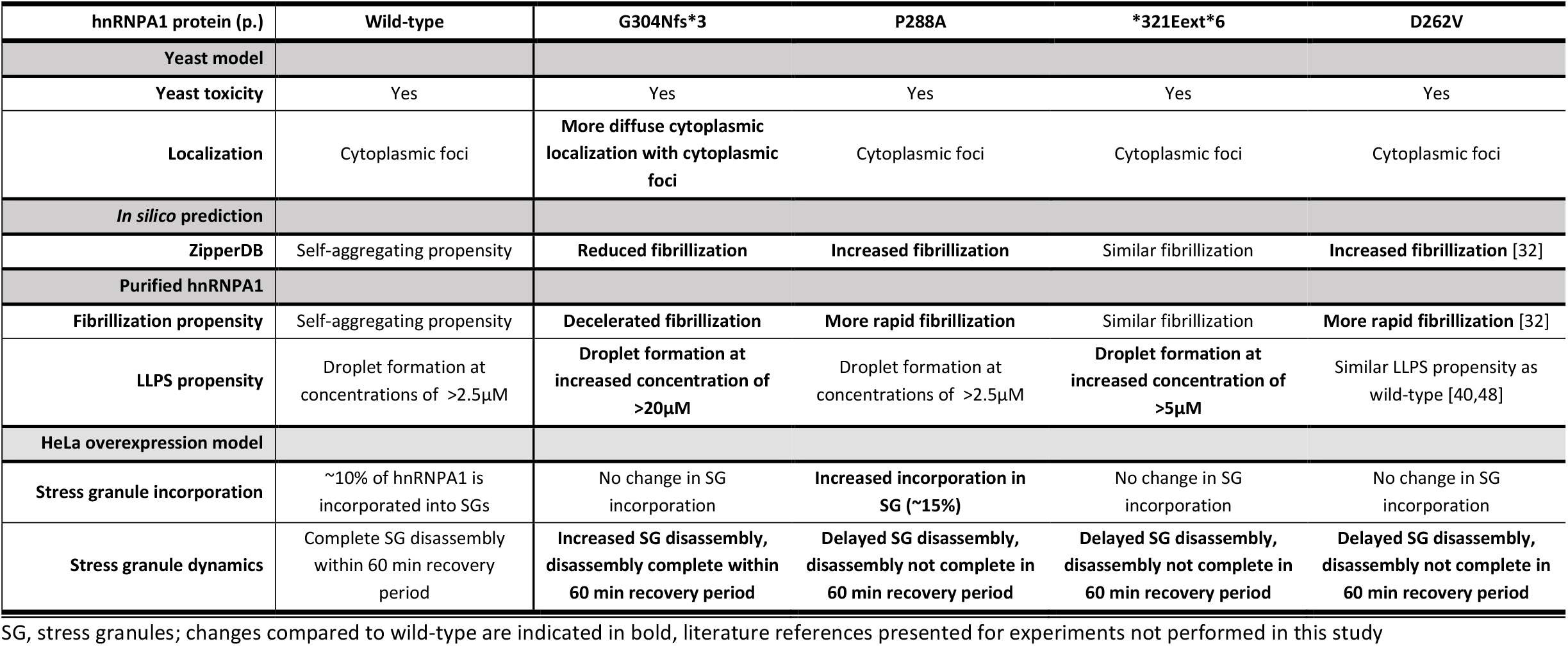
Summary of the molecular features of hnRNPA1 mutants.

| hnRNPA1 protein (p.) | Wild-type | G304Nfs*3 | P288A | *321Eext*6 | D262V |
| --- | --- | --- | --- | --- | --- |
| Yeast model |  |  |  |  |  |
| Yeast toxicity | Yes | Yes | Yes | Yes | Yes |
| Localization | Cytoplasmic foci | More diffuse cytoplasmic localization with cytoplasmic foci | Cytoplasmic foci | Cytoplasmic foci | Cytoplasmic foci |
| *In silico* prediction |  |  |  |  |  |
| ZipperDB | Self-aggregating propensity | Reduced fibrillization | Increased fibrillization | Similar fibrillization | Increased fibrillization [32] |
| Purified hnRNPA1 |  |  |  |  |  |
| Fibrillization propensity | Self-aggregating propensity | Decelerated fibrillization | More rapid fibrillization | Similar fibrillization | More rapid fibrillization [32] |
| LLPS propensity | Droplet formation at concentrations of >2.5μM | Droplet formation at increased concentration of >20μM | Droplet formation at concentrations of >2.5μM | Droplet formation at increased concentration of >5μM | Similar LLPS propensity as wild-type [40,48] |
| HeLa overexpression model |  |  |  |  |  |
| Stress granule incorporation | ~10% of hnRNPA1 is incorporated into SGs | No change in SG incorporation | Increased incorporation in SG (~15%) | No change in SG incorporation | No change in SG incorporation |
| Stress granule dynamics | Complete SG disassembly within 60 min recovery period | Increased SG disassembly, disassembly complete within 60 min recovery period | Delayed SG disassembly, disassembly not complete in 60 min recovery period | Delayed SG disassembly, disassembly not complete in 60 min recovery period | Delayed SG disassembly, disassembly not complete in 60 min recovery period |

SG, stress granules; changes compared to wild-type are indicated in bold, literature references presented for experiments not performed in this study

ZipperDB also revealed that the G304Nfs∗3 mutation deletes several potent steric zippers from the hnRNPA1 PrLD, which would likely reduce the multivalency of the PrLD and reduce fibrillization. We confirmed this prediction experimentally as the hnRNPA1^G304Nfs∗3^ variant exhibited reduced fibrillization *in vitro*. Finally, ZipperDB predicted that the ∗321Eext∗6 mutation does not significantly alter the steric zipper landscape of hnRNPA1. Accordingly, hnRNPA1^∗321Eext∗6^ exhibited similar fibrillization propensity as wild-type hnRNPA1.

In addition to forming stable fibrils, the low complexity PrLD can interact with each other and mediate LLPS, a process that contributes to SG assembly [48,40]. In this study, we observed that the G304Nfs∗3 variant also showed reduced propensity for LLPS, whereas the P288A and ∗321Eext∗6 variants were more similar to wild-type hnRNPA1. Although liquid condensates and stable fibrils are distinct material states, LLPS and fibrilization can conspire to promote pathological protein aggregation. First, mutations that lead to increased pathogenic fibrillization also cause phase-separated droplets to stabilize over time, promoting formation of persistent SGs [48]. Second, mutations in the PrLD that increase LLPS can stimulate fibrillization, as prolonged existence in a locally concentrated phase-separated environment drives fibril formation [32,48].

Under stress conditions, wild-type hnRNPA1 partitions into SGs, only to disperse again when the stress is relieved. We have observed that *HNRNPA1* mutations can change the fibrillization propensity of the protein, and can affect SG dynamics leading to altered SG assembly and/or disassembly. The highly fibril-prone P288A mutation also led to increased hnRNPA1 accumulation in SGs when expressed in stressed cells, and this mutation prolonged SG disassembly. Conversely, the G304Nfs∗3 variant shows a reduced tendency for fibrillization and facilitated more rapid SG disassembly. For the ∗321Eext∗6 variant, we observed fibrillization propensity similar to wild-type hnRNPA1 in combination with delayed SG disassembly. It remains unclear how the ∗321Eext∗6 mutant would slow SG disassembly without affecting fibril formation and LLPS, although perhaps the C-terminal -ELGNKA extension provides a short-linear motif that enables interactions that increase SG stability *in vivo*.

Intracellular inclusions of aggregated RBPs are one of the hallmarks of ALS/FTD. However, different hypotheses have been proposed about exactly how mutations in these proteins would cause neurodegeneration. On the one hand, pathological aggregates can cause a toxic gain-of-function. On the other hand, mutations can lead to loss of normal function [41]. Given its key roles in cellular functioning, loss of hnRNPA1 function could affect splicing, translation, and transcription of many targets. Furthermore, loss of hnRNPA1 function can alter miRNA biogenesis, and the implicated miRNAs in turn can disturb expression of their target transcripts. In addition, disruptions to normal SG formation can recruit other, wild-type PrLD-containing proteins to SGs, also affecting their normal activities [48,40].

Our results show that hnRNPA1 mutant proteins have different effects on the dynamics of SGs and the aggregation of RNA-binding proteins, whilst still causing phenotypes within the known spectrum of disease (Table 1, 2). All three mutant hnRNPA1 proteins altered SG dynamics: P288A and ∗321Eext∗6 slow SG disassembly, whereas the G304Nfs∗3 mutant accelerated SG disassembly. These findings suggest that rather than a universal mechanism defined by aggregates, there are multiple mechanisms at play (Table 2). However, which critical functions are most affected and how they impact on physiological pathways remains to be investigated. Further research into the differences in normal functions affected by the mutations might be able to explain some of the phenotypic variance observed for *HNRNPA1* mutations.

Why mutations in hnRNPA1, a ubiquitously expressed protein, have such a tissue-selective effect remains unknown. In the case of motor neurons, the extreme cell polarity and the great distance between the nucleus and cytoplasmic site of action may accentuate detrimental effects of altered nuclear-cytoplasmic shuttling of hnRNPA1, as with the P288A variant. This mutation is reminiscent of FUS PY-NLS mutations (e.g. P525L), which presents with persistent cytoplasmic mislocalization of FUS [11]. Like prions, self-templating conformers that can be transmitted between individuals, PrLD-containing proteins associated with cytoplasmic inclusions can show spreading to neighboring cells. For example, TDP-43 may be bi-directionally transmitted across synaptic terminals [12]. We cannot rule out that intercellular transmission contributes to the pathology caused by hnRNPA1 mutations. The identification of additional mutations in hnRNPA1 and other RNA- and DNA-binding proteins in degenerative disorders of motor neurons and muscle will help to better understand the underlying pathomechanisms. Based on our results, there clearly is not one unifying mechanism at play but rather a spectrum of various disturbances of fibrillization propensity, LLPS propensity, and SG dynamics.

## Data Availability

Sequence datasets of the six families are not available due to privacy concerns.

## ACKNOWLEDGEMENTS

We thank all the patients and family members for their participation in this study. Several authors of this publication are member of the European Reference Network for Rare Neuromuscular Diseases (ERN EURO-NMD) and of the Undiagnosed Diseases Network (UDN).

## SUPPLEMENTARY INFORMATION

**Fig. A1.**
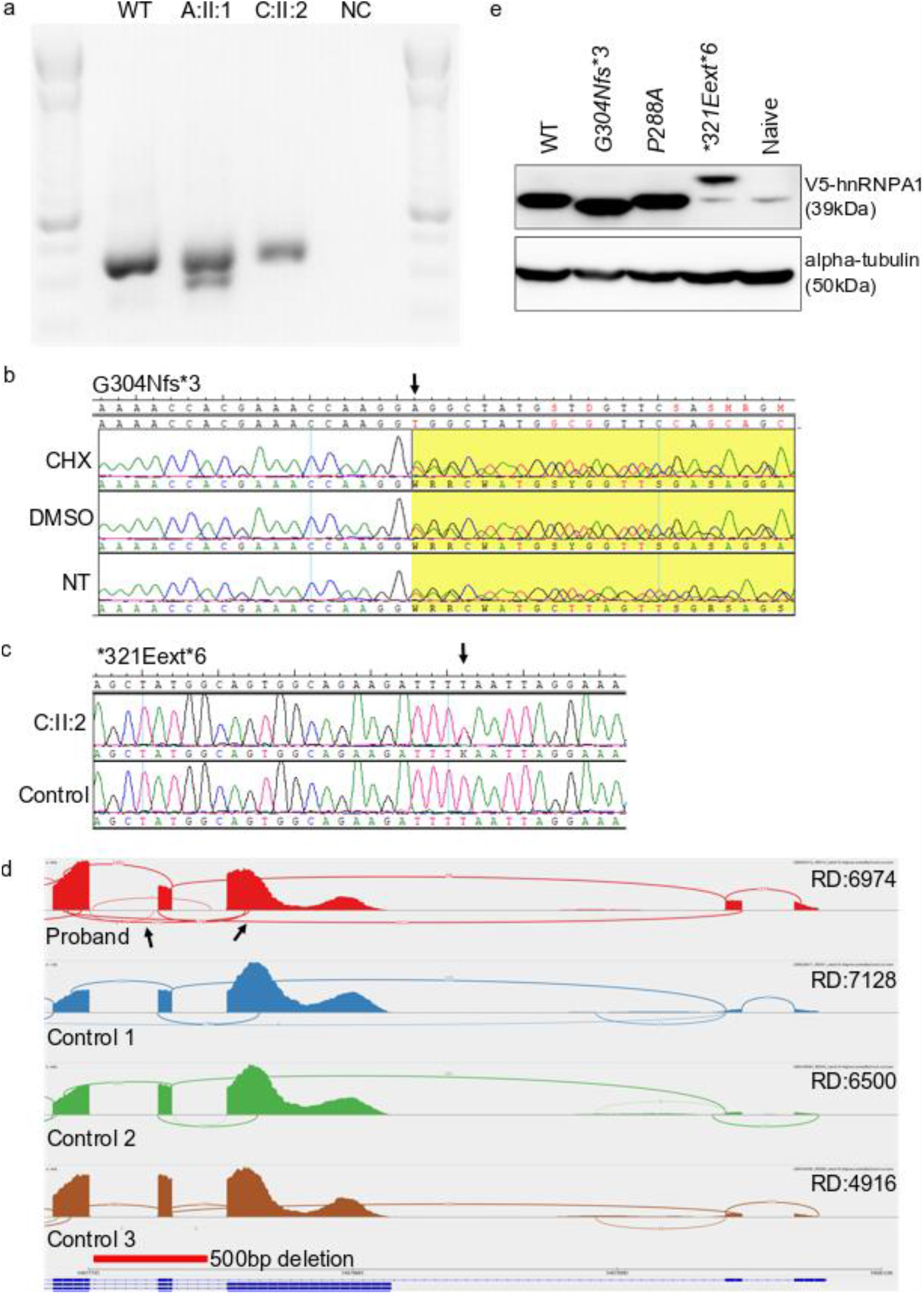
Validation of HNRNPA1 mutations on mRNA level and splicing, as well as protein expression in HeLa overexpression model. **a**. cDNA PCR results on agarose gel showing presence of shorter transcript in family A in addition to the wild-type band. Second transcript for family C is not visible on gel. **b**. cDNA sequencing at mutation site for the G304fs∗3 mutation in family A, showing the mutation in heterozygous state in all three conditions (cycloheximide CHX, vehicle control DMSO and non-treated NT), thus not subjected to nonsense-mediated decay. **c**. Sanger sequencing of cDNA of patient C:II:2 and a healthy control, showing the presence of the ∗321Eext∗6 mutation on mRNA level **d**. Sashimi plot demonstrating abnormal skipping of exon 9 NM_002136.4 (indicated by arrows) in D:II:1 proband (top row) when compared to control samples. Read depth (RD) listed per sample. The 500bp deletion shown as relative to the genomic location and NM_002136.4 transcript **e**. Western blot results of transfected HeLa cells with HNRNPA1-constructs showing bands of the expected size for all mutants. Wild-type HNRNPA1-V5 results in a band of approximately 39kDa, as does the P288A mutant. Whereas the G304Nfs∗3 and the ∗321Eext∗6 mutations result in a slightly smaller and slightly larger protein respectively.

**Fig. A2.**
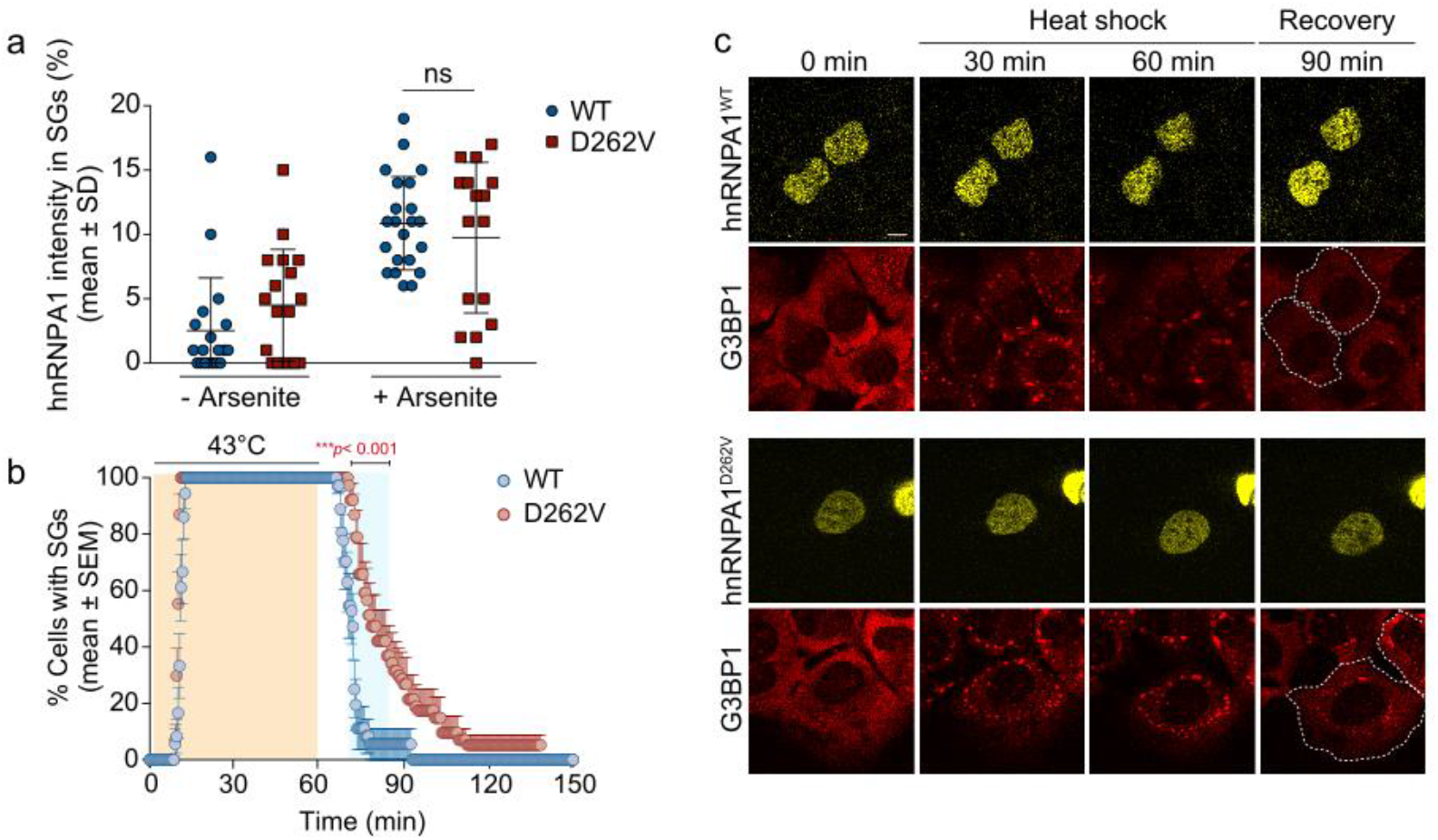
Localization and stress granule dynamics of the hnRNPA1 D262V mutant. **a**. HeLa cells were transiently transfected with WT or mutant EGFP-tagged hnRNPA1 and subjected to arsenite stress (0.5 mM sodium arsenite, 30 min). Cells were fixed and stained with DAPI, eIF3η, and G3BP, and the intensity of hnRNPA1 signal in stress granules in each cell was measured. An interleaved scatter plot with individual data points is shown. Error bars represent mean ± SD (n = 18 and 22 cells for hnRNPA1 and hnRNPA1^D262V^, respectively). ns, not significant by two-way ANOVA with Sidak’s multiple comparisons test. Scale bar, 10 μm. **b**,**c**. U2OS cells expressing tdTomato-tagged endogenous G3BP1 were transiently transfected with WT or mutant EYFP-tagged hnRNPA1 and subjected to heat shock (43 °C, 60 min; orange shading) and allowed to recover at 37 °C for 2 h. White dotted lines delineate hnRNPA1-positive cells. Line graph in **c** represents the percentage of cells with visible tdTomato-G3BP1 puncta over time (n = 18 and 19 videos for hnRNPA1 and hnRNPA1^D262V^, respectively). Blue shaded area indicates time points at which D262V mutant was statistically significantly different from WT. ∗∗∗p < 0.001, by two-way ANOVA with Sidak’s multiple comparisons test. Scale bar, 10 μm.

